# SWPER Global: A survey-based women’s empowerment index expanded from Africa to all low- and middle-income countries

**DOI:** 10.1101/2020.07.31.20166223

**Authors:** Fernanda Ewerling, Anita Raj, Cesar G. Victora, Franciele Hellwig, Carolina V. N. Coll, Aluisio J. D. Barros

**Affiliations:** International Center for Equity in Health, Federal University of Pelotas, Pelotas, Brazil; Postgraduate Program in Epidemiology, Federal University of Pelotas, Pelotas, Brazil; Center on Gender Equity and Health, University of California San Diego, US

## Abstract

**Introduction:** In 2017, a survey-based women’s empowerment index (SWPER) was proposed for African countries, including three domains: social independence, decision making and attitude to violence. External validity and predictive value of the SWPER has been demonstrated in terms of coverage of maternal and child interventions and use of modern contraception. To determine its value for global monitoring, we explored the applicability of the SWPER in national health surveys from low- and middle-income countries (LMICs) in other world regions.

**Methods:** We used data from the latest Demographic and Health Survey for 62 LMICs since 2000. 14 pre-selected questions (items) were considered during the validation process. Content adaptations included the exclusion of women’s working status and recategorization of the decision-making related items. We compared the loading patterns obtained from principal components analysis performed for each country separately with those obtained in a pooled dataset with all countries combined. Country rankings based on the score of each SWPER domain were correlated with their rankings in the Gender Development Index (GDI) and the Gender Inequality Index (GII) for external validation.

**Results:** Consistency regarding item loadings for the three SWPER empowerment domains was observed for most countries. Correlations between the scores generated for each country and global score obtained from the combined data were 0.89 or higher for all countries. Correlations between the country rankings according to SWPER and GDI were, respectively, 0.74, 0.72 and 0.67 for social independence, decision-making, and attitude to violence domains. The correlations were equal to 0.81, 0.67, and 0.44, respectively, with GII.

**Conclusion:** The indicator we propose, named SWPER Global, is a suitable common measure of women’s empowerment for LMICs, addressing the need for a single consistent survey-based indicator of women’s empowerment that allows for tracking of progress over time and across countries at the individual and country levels.

**Summary:** *What is already known?:*

- Although survey-based women’s empowerment indicators have been used in the literature, until 2017 there was no indicator proposed for use in a large set of countries that would be comparable between and within countries.
- In 2017, we proposed the Survey-based Women’s emPowERment indicator (SWPER, pronounced as “super”), to be used in African countries, that encompasses three well recognized domains of women’s empowerment (attitude to violence, social independence and decision making).
- The external validity and predictive value of the SWPER has been demonstrated in terms of coverage of maternal and child interventions and use of modern contraception.
- Validation of the index was restricted to African countries, and a common measure to allow comparisons across low and middle-income countries (LMICs) from all world regions was still lacking.

*What are the new findings?:*

- We show that the SWPER Global may serve as a valid common measure of women’s empowerment among LMICs, as consistent patterns were obtained for most countries and world regions.
- The SWPER Global index addresses the need for a single cross-cultural standardized survey-based indicator of women’s empowerment in the context of LMICs that enables comparability between countries and over time and subgroup analyses, extending previously proposed indicators such as the Gender Development Index which is limited to the country-level

*What do the new findings imply?:*

- The SWPER Global index enables the study of how women’s empowerment is linked to developmental and health outcomes, allowing for broad comparisons across countries and world regions.
- As a comprehensive cross-cultural standard tool, it also contributes to the monitoring and accountability of country progress over time in advancing gender equality and women’s empowerment.
- The new tool may help target and prioritize policy and advocacy efforts toward SDG 5 (achieve gender equality and empower all women and girls) at the regional and country levels.

## Introduction

Empowerment is a complex and multidimensional construct, often defined both as a process and an outcome, by which individuals gain power over their lives and decisions (1). To be empowered, women must not only have key assets (such as education and health) and access to opportunities (such as employment), but also have the agency (i.e., perceived and actual self-efficacy and decision-making control) to move from making planned choices to achieving one’s self-determined goals (2,3). At the individual level, empowerment involves women utilizing their assets, opportunities, and agency for making purposive choices and engaging in behaviors to alter life circumstances. This may include engagement in positive behaviors and action towards health and economic development (4–6). The Millennium Development Goals raised the women’s empowerment agenda in the recent past, by recognizing its importance for health as well as development, yet gender inequalities persist (7,8). Women’s Human Development Index is, on average, 6% lower than that of men, with the widest gaps observed in the poorest countries. Much of the gap is due to women’s lower educational attainment, economic participation and income (7,9). In 2015, the fifth Sustainable Development Goal (SDG) called for “achievement of gender equality and the empowerment of all women and girls” as a vital goal for accelerating sustainable development. However, there is no consensus on how to measure women’s empowerment and an internationally standardized l indicator is still unavailable, which precludes accountability in low-and middle-income countries (LMICs) (10).

In the recent years, some measures that capture gender inequalities in key socioeconomic and health indicators (e.g., Gender Development Index, Global Gender Gap Index (7)), gender discrimination and gender-related risks for women such as maternal mortality or adolescent motherhood (e.g., Social Institutions and Gender Index (11), and the Gender Inequality Index (8)), or women’s general wellbeing including safety from violence such as the Women, Peace and Security Index (12), have been proposed in the literature. However, most of these indices rely on national-level aggregate data and cannot be disaggregated by region or population subgroups, limiting within-country comparisons. Consideration of subgroups is very important to account for intersectionality, as it is known that socially marginalized women, including those who are poorer, rural, less educated or reside in fragile states, face greater risks to their health, well-being and even survival (13,14). Intersectionality is often ignored (15). A few individual-level measures were also proposed (10,16,17), but most of these tools were specifically designed for a given country or region, or a particular sector such as the Women’s Empowerment in Agriculture Index, which has a particular focus on women who are working in agriculture (18,19).

In the context of LMICs, national Demographic and Health Surveys (DHS) are a reliable source of individual level information on socioeconomic characteristics, health, and development indicators and, since 1999, have been incorporating questions on women’s empowerment that potentially allows for comparisons within and between countries using an intersectional lens.

In 2017, a survey-based women’s empowerment index (SWPER) was developed and validated using DHS data from 34 African countries (10). SWPER captures three empowerment domains indicative of assets and agency:

1. Social independence: mainly composed by preconditions that enable women to achieve their goals (schooling attainment, access to information, age at pivotal life events and spousal asset differentials and access to information).
2. Decision-making: the extent of the woman’s participation is household decisions, which may also be considered a measure of instrumental agency.
3. Attitudes to violence: closely related to the concept of intrinsic agency, as a proxy for the woman’s incorporation of gender norms related to wife-beating acceptability.

The development of the SWPER used a similar conceptual framework further recently proposed by Miedema *et al* (16), which describes three domains of empowerment: enabling conditions, instrumental agency and intrinsic agency. Enabling conditions are considered preconditions that allow women to gain more power (20), and these correspond to the asset and opportunity constructs commonly focused upon in development economists’ definitions of empowerment (2,4). Instrumental agency is the woman’s ability to make choices in the household, at family-level (16). Intrinsic agency – or power within – is the process by which one develops a critical consciousness of one’s own aspirations, capabilities, and rights (16,21,22), and can be viewed as an asset and opportunity regarding safety of the environment or as a proxy for agency as safety would allow for greater ability to act on choice (16).

The SWPER uses individual-level data, allowing for assessing associations between empowerment and several health interventions and outcomes (10). The SWPER also allows within-country and between-country comparisons, as well as time trend analyses. The items used to calculate the SWPER are available for over 60 countries with a DHS. The external validity and predictive value of the African SWPER has been demonstrated in terms of coverage of maternal and child interventions and use of modern contraception (10). The SWPER attracted interest from the academic community and international agencies. In July 2018, a workshop with experts on women’s empowerment was held in Washington, DC, co-organized by PAHO and the Countdown to 2030 (23,24) to allow a comprehensive debate on definitions of women’s empowerment and the expansion of SWPER beyond Sub-Saharan Africa, especially in respect to the Latin American context (see Panel 1 for details). Feedback from the workshop prompted us to develop and test a global version of the SWPER that would allow its use in all LMICs.

In July 2018, an expert workshop was held in Washington, DC, bringing together a panel of experts with the aim of discussing how to best adapt the SWPER for use in Latin America and other world regions (https://equidade.org/news/123/expert-group-workshop-on-women-39-s-empowerment-in-the-lac-region). The workshop was co-organized by PAHO/WHO and the Countdown to 2030 and counted with the participation of over 20 experts from multilateral organizations and universities (participants will be listed in the acknowledgements upon their agreement).

A series of suggestions for improving the indicator were made, notably:

1. Exclude the woman’s working status item, which was considered too simplistic to indicate whether or not work was empowering women, given that women may work because they are forced to work due to circumstance, and may not even be paid for this work;
2. For the questions on who decides on health care utilization and household expenses, give equal weight to joint decisions (with the partner or another person) and woman’s deciding alone;
3. Add items related to sexual and reproductive autonomy, decision-making on the use of the woman’s income, ownership of house or land; and access to technology, such as mobile phones; and
4. Include unpartnered women to allow the assessment of empowerment in this group.

The currently proposed version of the SWPER Global incorporates the first two recommendations. Given the constraints of the data currently available in surveys and the need for a readily available measure of women’s empowerment, the latter two recommendations were not incorporated at this time. We acknowledge these limitations and commit to continue our efforts to refine the indicator in the future.

**Panel 1. Summary of the recommendations from the expert workshop held in Washington, DC, to discuss the adaptation of the SWPER for LMICs in Latin America and other world regions**.

## Methods

We used data from the latest available DHS since 2000 for all available countries. For Mozambique, data from the 2011 survey was used because the 2015 survey lacked information on partner’s age and education. Six countries with a DHS were dropped because not all necessary items were available (Colombia, Turkey, Vietnam, Jordan, Yemen and Congo Brazzaville). Thus, 62 countries – 34 of which were already included in the SWPER for Africa (10) – were included in the analyses (Table 1), with a total sample of 662,835 partnered women.

**Table 1.**
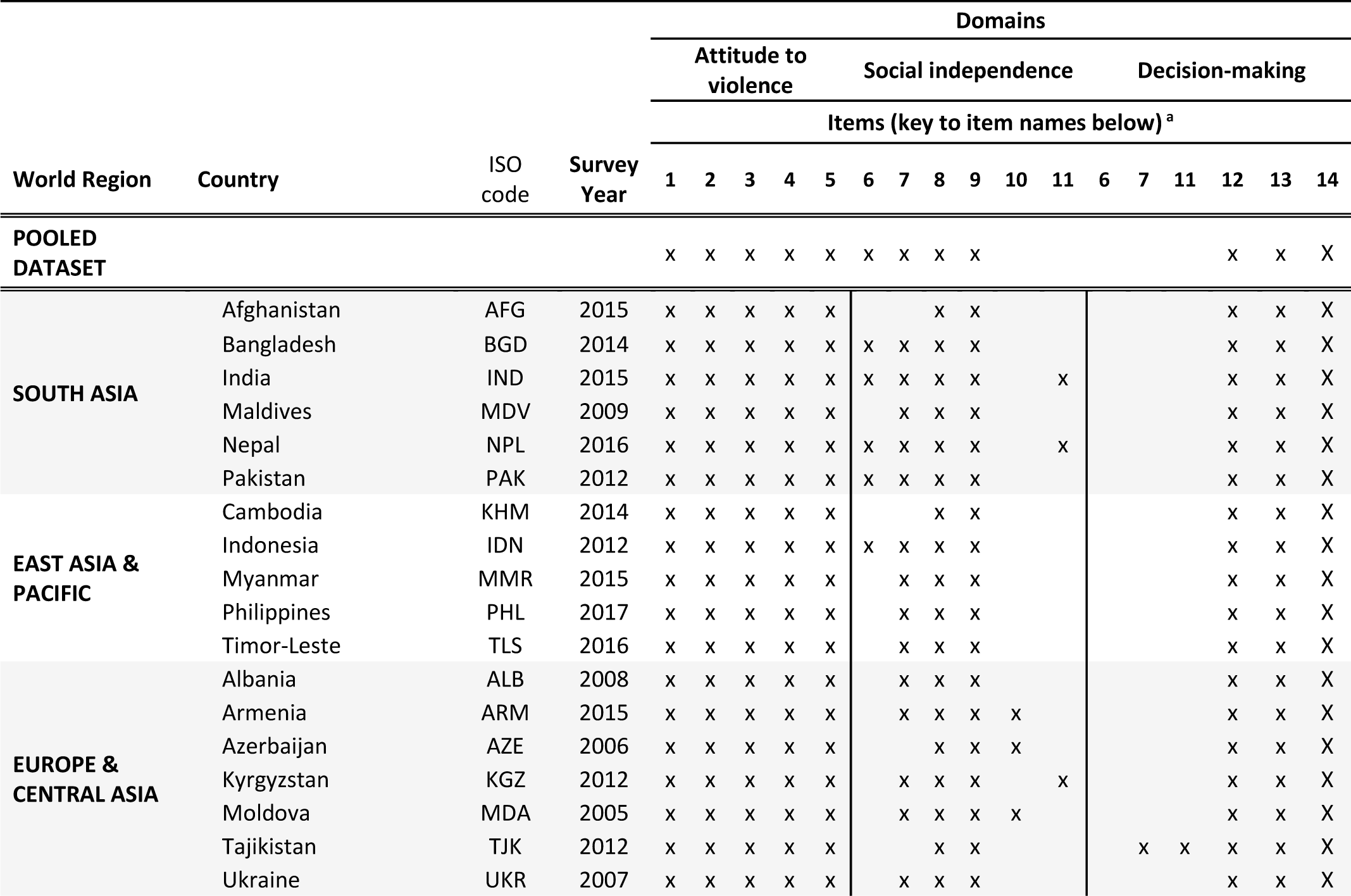
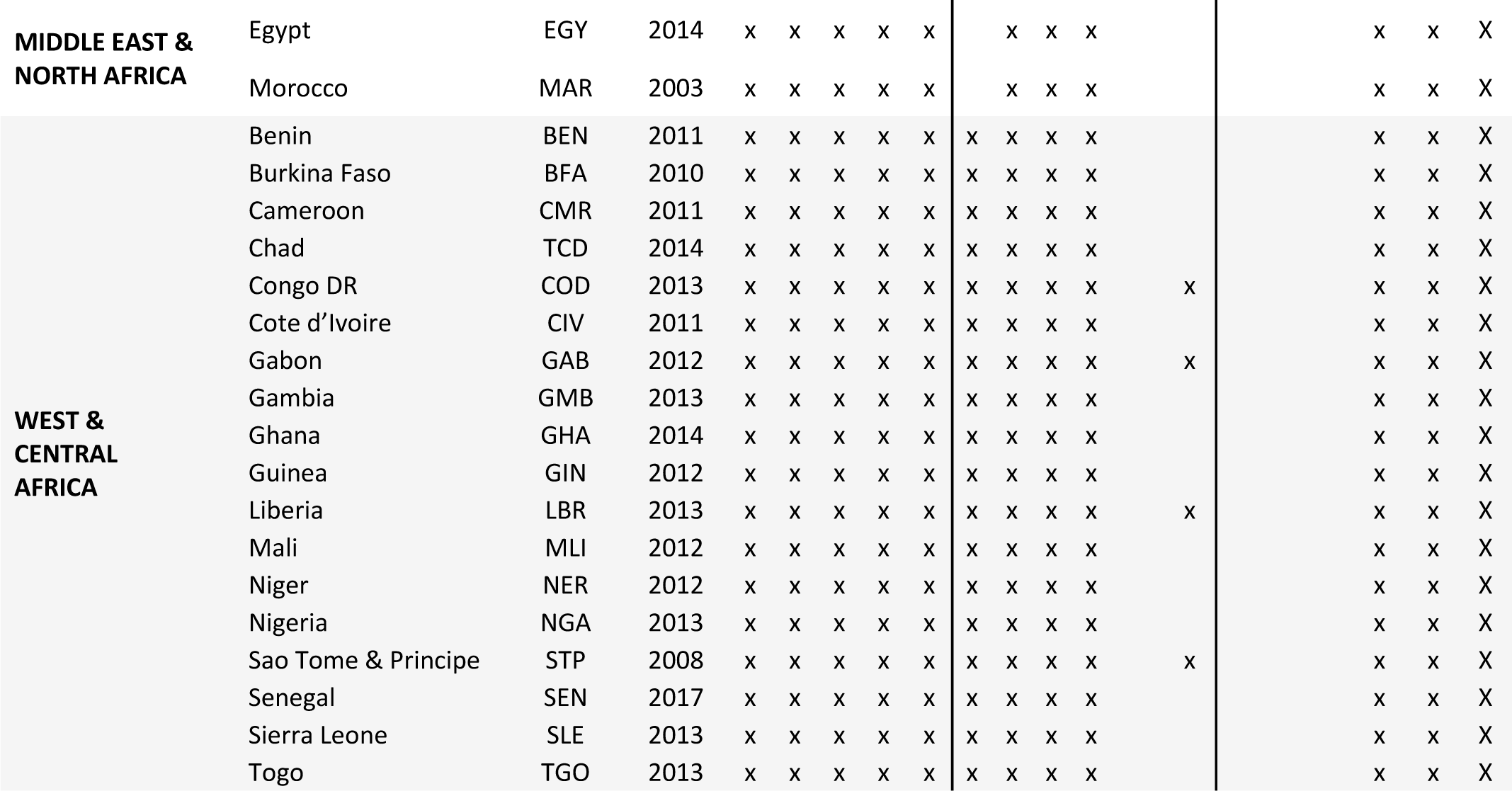
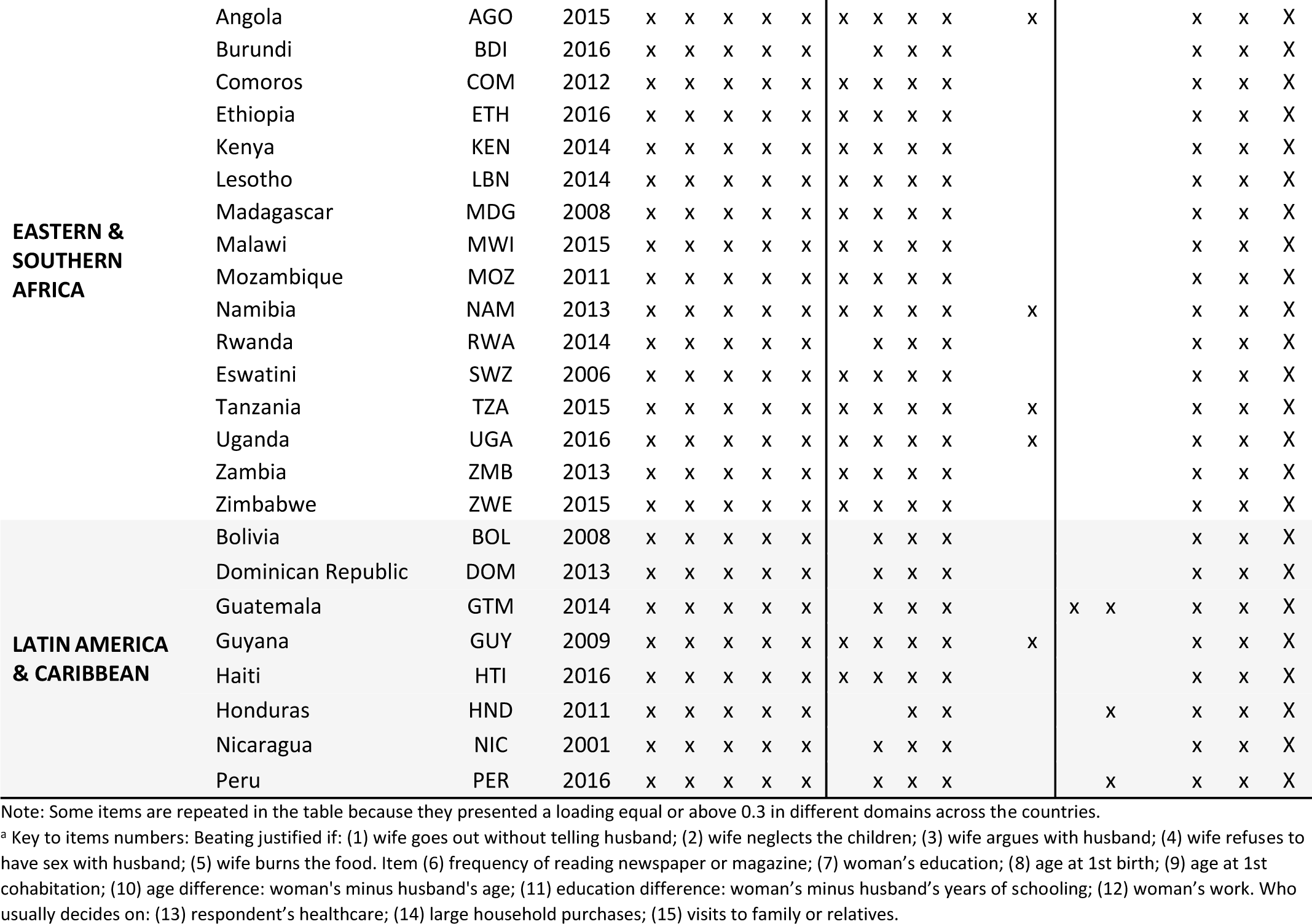
Composition patterns of the items that compose the SWPER domains with loadings’ equal or above 0.3.

### Content validity

The SWPER Global was developed using similar methods to what was done for the SWPER for Africa. In that case, we used 15 items available in DHS surveys (see Table 2) and the scores were derived through principal components analyses (PCA) using surveys from 34 African countries (10). We obtained an indicator with three domains: (1) attitude to violence, based on five questions asking the women’s opinion on whether a husband beating the wife is justified in specific situations; (2) social independence, comprising access to information, educational attainment, age at marriage and first child, and differences in age and education to the cohabiting partner; and (3) decision-making, based on questions related to who makes decisions in the household (in regard to the respondent’s health care, large purchases and to visits to family or relatives) and to the women’s work.

**Table 2.**
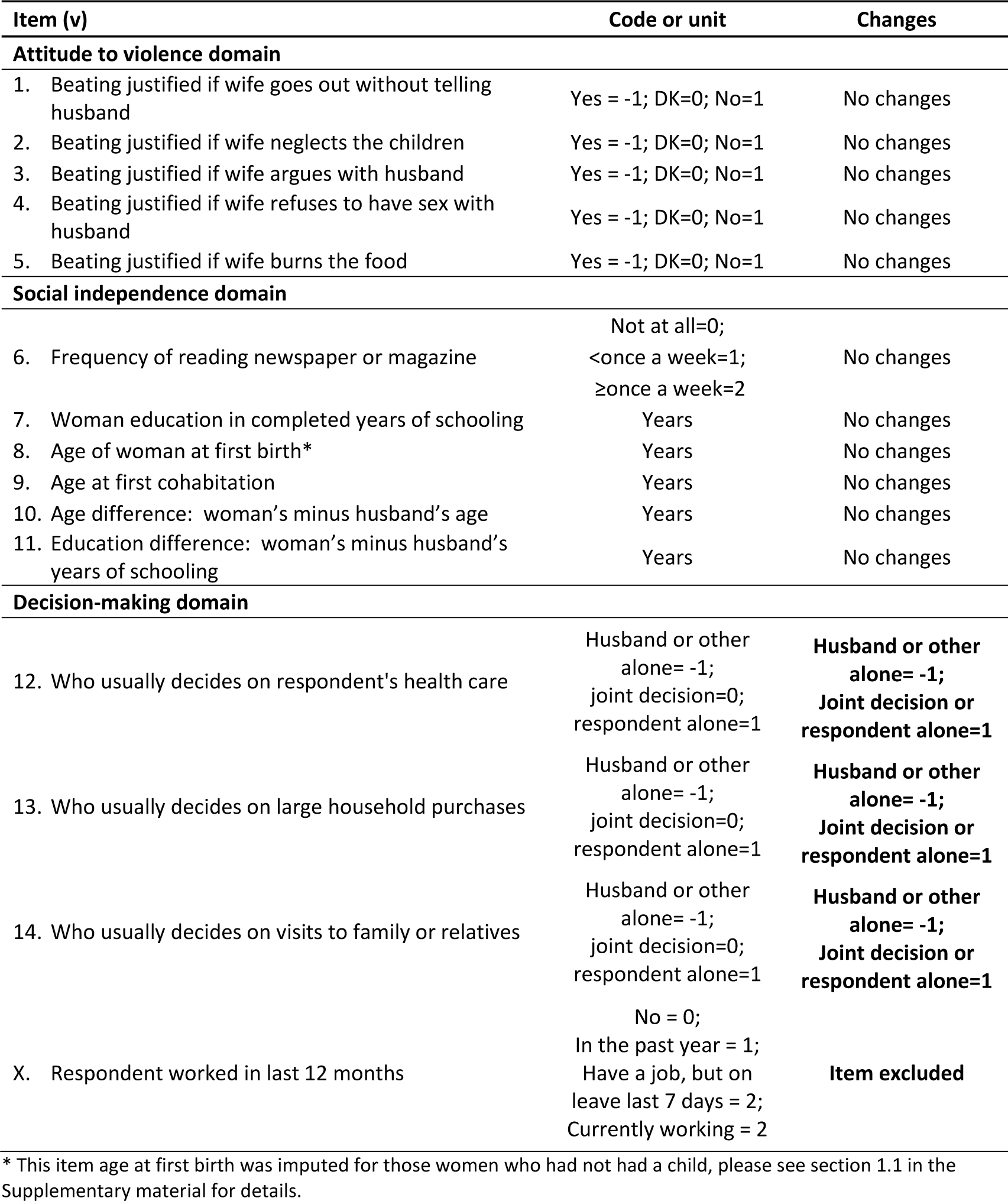
Items used in each domain of the African-oriented survey-based women’s empowerment (SWPER) index and the changes made in the global version of the index, according to gender experts’ recommendations.

Considering the expert workshop recommendations (Panel 1), we excluded the item that indicates whether the woman worked in the last year and changed the categorization of the decision-making related variables, so that equal weights were given for joint decisions (with the partner or another person) and woman’s sole decision (See Table 2 for details).

### Construct validity

Following a similar methodology used for Africa (10), we used PCA to identify the empowerment domains and estimate the items loadings for each of the 62 countries separately, after applying varimax rotation. We then applied the same strategy to a pooled dataset combining all countries, to derive a single indicator. The patterns emerged were compared in terms of items with loadings of 0.3 or higher in each empowerment domain across countries to check for consistency, and Pearson correlation coefficients between the scores derived for each individual country and global scores derived from the combined data for each empowerment domain of the indicator were estimated. By doing so, we aimed to evaluate whether these two approaches would present consistent estimates of individual empowerment levels.

### External validity

To assess the external validity of the SWPER for this extended set of LMICs, Spearman correlation coefficients were calculated between the resulting pooled score and two widely used indices: the Gender Development Index (GDI) and the Gender Inequality Index (GII) (8). In this case, Spearman correlation was used because we were interested in the correlation between the ranking of the countries using these different measures, rather than the correlation between the scores.

The equations to calculate the SWPER Global are provided in the Supplementary materials with a step-by-step explanation. To allow researchers to choose the reference population and standardize the SWPER scores according to the world region they consider more suitable for a given study, we also calculated the means and standard deviations of the SWPER Global domains for each world region additionally to the scores estimated for all countries together (Table S2 in the web appendix). With that, a researcher interested in India, for example, could choose South Asia or the SWPER Global (all countries together) as reference population to standardize the SWPER scores. In the Results section below, we used the global mean for LMICs and standard deviation to standardize the scores. Based on the distribution of scores, we also proposed a categorization of the SWPER domains in three groups: low, medium and high empowerment level. For the social independence domain, for which the distribution to a normal curve, the scores were divided into terciles. In contrast, the attitude to violence and decision-making domains present multimodal distributions, which were considered to define the cut-offs (see Figure S1 and Table S3 in the web appendix).

All analyses were performed using the statistical software Stata (StataCorp. 2017. Stata Statistical Software: Release 15.1. College Station, TX: StataCorp LLC). The calculation of average scores at country level took into account the surveys’ sample design. DHS are public sources of information and ethical approval has already been obtained from each country by the time of the survey conduction.

## Results

The same three domains identified in the SWPER for Africa were observed in the global analyses: attitude to violence, social independence and decision making. The cross-country consistency among items composing each domain of the SWPER Global can be assessed through Table 1, where we present items with a loading of 0.3 or more (henceforth referred to as high loadings). All countries presented the exact same pattern of items identified in the attitude to violence domain, which included all the questions related to the woman’s opinion on whether a husband is justified in beating the wife in five different circumstances. Items composing the decision-making domain was also consistent across countries with higher loadings for the three questions related to the women’s involvement in household decisions, with only four countries presenting high-loading items that were not related to participation in household decisions. Three of them were from Latin America (Honduras and Peru, that also included woman’s education; and Guatemala, that included education and frequency of reading newspaper or magazine) and one from Europe & Central Asia (Tajikistan, that also included woman’s education and education difference between the woman and her husband). For the social independence domain, a greater variability in the patterns of items identified was observed. Countries from Africa and South Asia were the most stable in terms of the patterns for this domain (composed by the frequency of reading newspaper/magazine, women’s education, age the first birth, age at first cohabitation, and age and education differentials in relation to the partner). In the other four regions of the world, most countries presented low loadings for the frequency of reading newspapers or magazines. Education showed low loadings in only five countries (Azerbaijan, Tajikistan, Afghanistan, Cambodia and Honduras). The items related to pivotal events (age at first birth and at first cohabitation) presented high loadings in all countries. Age differentials to the partner had high loadings in three Europe and Central Asia countries (Armenia, Azerbaijan and Moldova) and education differentials to the partner in 12 out of the 62 countries, but without a clear pattern.

In Table 3 we present the correlations between the three SWPER domain scores calculated separately using principal component analyses in each country, and the global scores calculated using the pooled data. Cells are colored from yellow to dark green cells showing the strength of the correlations. Even though the item patterns presented in Table 1 were not entirely consistent for some countries (i.e. all items presenting loading ≥0.3 in the same empowerment domain), the correlations between the country-specific and the SWPER Global scores were very high, equaling 0.89 or more. All countries presented correlations greater than 0.91 for all three SWPER domains, except for Gabon and Liberia where the correlations for social independence were 0.89 and 0.90, respectively.

**Table 3.** Pearson correlation between the country-specific women’s empowerment measure and the SWPER global index for each domain. Cells are colored from yellow (r<0.900), light green (r=0.900 to <0.950), medium green (0.950 to <0.990), dark green (≥ 0.990).

| World region | Country | Year | Pearson's correlation (r) |  |  |
| --- | --- | --- | --- | --- | --- |
|  |  |  | Attitude to violence | Social Independence | Decision Making |
| SOUTH ASIA | Afghanistan | 2015 | 0.9923 | 0.9545 | 0.982 |
|  | Bangladesh | 2014 | 0.9968 | 0.9898 | 0.9936 |
|  | India | 2015 | 0.9983 | 0.9935 | 0.994 |
|  | Maldives | 2009 | 0.9976 | 0.9819 | 0.9687 |
|  | Nepal | 2016 | 0.9872 | 0.9896 | 0.9911 |
|  | Pakistan | 2012 | 0.9996 | 0.9966 | 0.995 |
| EAST ASIA & PACIFIC | Cambodia | 2014 | 0.9901 | 0.9776 | 0.9685 |
|  | Indonesia | 2012 | 0.9918 | 0.993 | 0.9928 |
|  | Myanmar | 2015 | 0.9864 | 0.993 | 0.9636 |
|  | Philippines | 2017 | 0.9753 | 0.9911 | 0.9862 |
|  | Timor-Leste | 2016 | 0.9942 | 0.9933 | 0.9813 |
| EUROPE & CENTRAL ASIA | Albania | 2008 | 0.9959 | 0.9807 | 0.9743 |
|  | Armenia | 2015 | 0.9846 | 0.9821 | 0.9848 |
|  | Azerbaijan | 2006 | 0.9884 | 0.9752 | 0.996 |
|  | Kyrgyzstan | 2012 | 0.9971 | 0.962 | 0.992 |
|  | Moldova | 2005 | 0.9908 | 0.9726 | 0.9579 |
|  | Tajikistan | 2012 | 0.9994 | 0.9807 | 0.9974 |
|  | Ukraine | 2007 | 0.967 | 0.989 | 0.9737 |
| MIDDLE EAST & NORTH AFRICA | Egypt | 2014 | 0.9978 | 0.9943 | 0.9955 |
|  | Morocco | 2003 | 0.99 | 0.9981 | 0.9955 |
| WEST & CENTRAL AFRICA | Benin | 2011 | 0.9977 | 0.9569 | 0.9968 |
|  | Burkina Faso | 2010 | 0.9991 | 0.9784 | 0.9758 |
|  | Cameroon | 2011 | 0.9992 | 0.9938 | 0.9965 |
|  | Chad | 2014 | 0.9934 | 0.9677 | 0.9778 |
|  | Congo DR | 2013 | 0.9964 | 0.9898 | 0.9972 |
|  | Cote d'Ivoire | 2011 | 0.9989 | 0.9526 | 0.9956 |
|  | Gabon | 2012 | 0.9973 | 0.8895 | 0.9901 |
|  | Gambia | 2013 | 0.9986 | 0.9858 | 0.9857 |
|  | Ghana | 2014 | 0.9986 | 0.9906 | 0.9979 |
|  | Guinea | 2012 | 0.9887 | 0.9639 | 0.995 |
|  | Liberia | 2013 | 0.999 | 0.8959 | 0.9944 |
|  | Mali | 2012 | 0.9966 | 0.9463 | 0.9943 |
|  | Niger | 2012 | 0.9991 | 0.9479 | 0.9944 |
|  | Nigeria | 2013 | 0.9988 | 0.9938 | 0.9971 |
|  | São Tome & Príncipe | 2008 | 0.9955 | 0.9649 | 0.9955 |
|  | Senegal | 2017 | 0.9988 | 0.9772 | 0.9821 |
|  | Sierra Leone | 2013 | 0.9985 | 0.9246 | 0.9956 |
|  | Togo | 2013 | 0.9983 | 0.984 | 0.9913 |
| **EASTERN &**<br>**SOUTHERN**<br>**AFRICA** | Angola | 2015 | 0.9981 | 0.9264 | 0.9786 |
|  | Burundi | 2016 | 0.9996 | 0.992 | 0.9936 |
|  | Comoros | 2012 | 0.9949 | 0.9876 | 0.9844 |
|  | Eswatini | 2006 | 0.989 | 0.9684 | 0.987 |
|  | Ethiopia | 2016 | 0.9991 | 0.9896 | 0.9976 |
|  | Kenya | 2014 | 0.9989 | 0.9962 | 0.9948 |
|  | Lesotho | 2014 | 0.9976 | 0.9945 | 0.9837 |
|  | Madagascar | 2008 | 0.9928 | 0.9906 | 0.9734 |
|  | Malawi | 2015 | 0.9962 | 0.9944 | 0.997 |
|  | Mozambique | 2011 | 0.9954 | 0.9895 | 0.9833 |
|  | Namibia | 2013 | 0.9966 | 0.9202 | 0.9932 |
|  | Rwanda | 2014 | 0.999 | 0.9992 | 0.9883 |
|  | Tanzania | 2015 | 0.9979 | 0.9754 | 0.9969 |
|  | Uganda | 2016 | 0.9995 | 0.9822 | 0.9835 |
|  | Zambia | 2013 | 0.9992 | 0.9899 | 0.9903 |
|  | Zimbabwe | 2015 | 0.999 | 0.9893 | 0.9918 |
| **LATIN AMERICA**<br>**& CARIBBEAN** | Bolivia | 2008 | 0.9897 | 0.9938 | 0.9683 |
|  | Dominican Republic | 2013 | 0.9414 | 0.9946 | 0.9883 |
|  | Guatemala | 2014 | 0.9877 | 0.9675 | 0.919 |
|  | Guyana | 2009 | 0.9967 | 0.9702 | 0.9927 |
|  | Haiti | 2016 | 0.9929 | 0.9853 | 0.9683 |
|  | Honduras | 2011 | 0.9927 | 0.9711 | 0.9242 |
|  | Nicaragua | 2001 | 0.9939 | 0.995 | 0.9897 |
|  | Peru | 2016 | 0.9643 | 0.9904 | 0.9372 |

Figure 1 shows scatter plots of the country ranks for the three SWPER domains against the country ranks for the GDI and the GII, providing evidence on the external validity of the SWPER. The correlation coefficients with the GDI (upper graphs) were 0.74, 0.72 and 0.67 for social independence, decision-making and attitude to violence, respectively. In the lower graphs, the SWPER rankings are plotted against the GII rankings. In this case, the Pearson correlations were 0.81, 0.67 and 0.44, respectively. Figure 1 shows that Latin America & Caribbean and Europe & Central Asia tend to present higher ranks in all empowerment domains while West & Central Africa, Eastern & Southern Africa and some countries from South Asia presented the lowest rankings.

**Figure 1.**
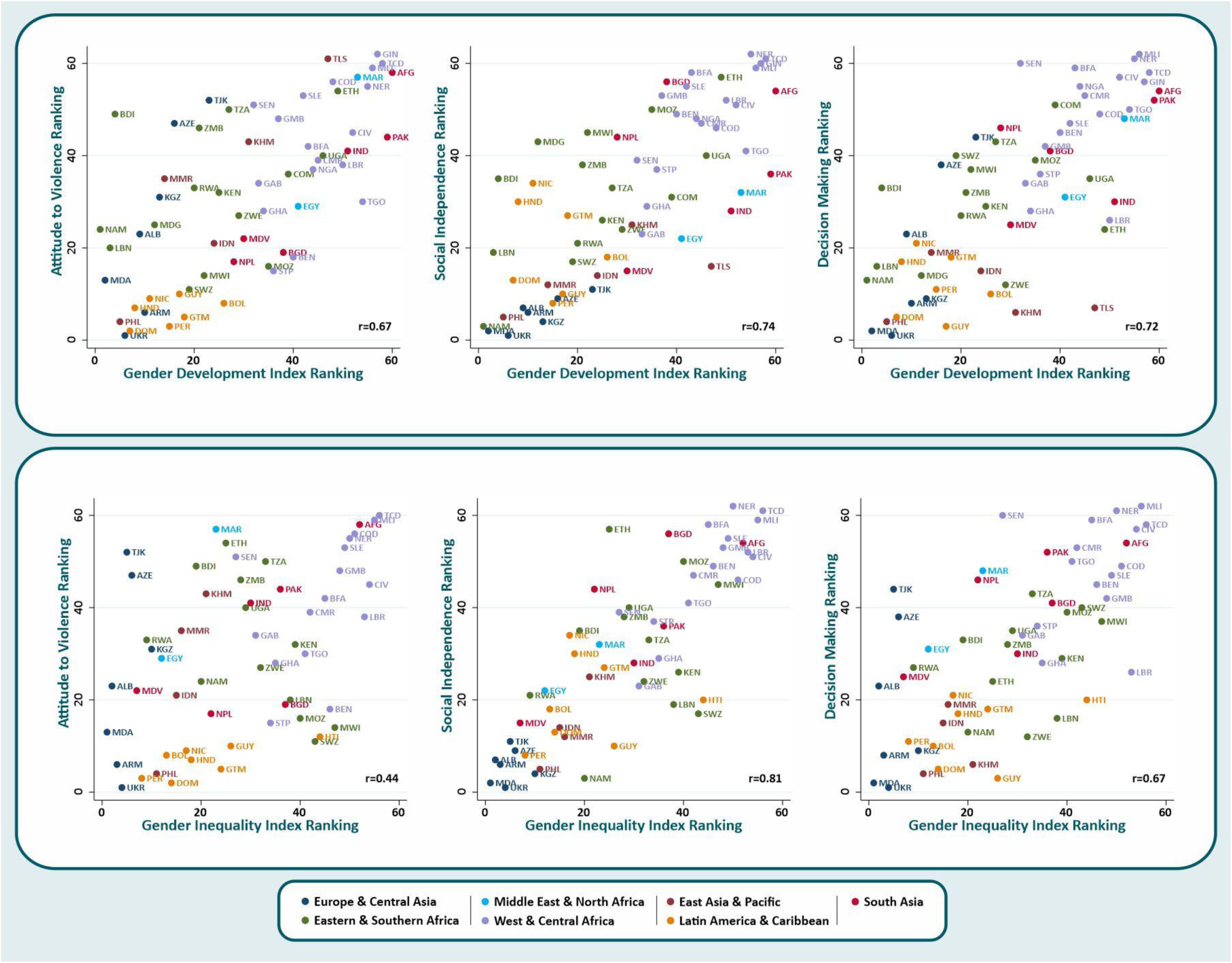
Scatter plot showing the SWPER domains ranking against the Gender Development Index and the Gender Inequality Index rankings with the Pearson correlation (r) indicated in the bottom right of each plot.A

Generally, Europe & Central Asia and Latin America & Caribbean show the higher mean scores in the three empowerment domains (Table 4). The African regions and South Asia presented the lower mean scores, with West & Central Africa showing the lowest average scores in all three empowerment domains. East Asia and Pacific presented high mean scores for social independence and decision making but scored below the average for attitude to violence.

**Table 4.** Mean and standard error of the SWPER global scores in each domain according to the world region.

| Region | Number of countries | Attitude to violence |  | Social Independence |  | Decision-making |  |
| --- | --- | --- | --- | --- | --- | --- | --- |
|  |  | Mean | Std. error | Mean | Std. error | Mean | Std. error |
| South Asia | 6 | 0.007 | 0.184 | -0.155 | 0.149 | 0.072 | 0.139 |
| East Asia & Pacific | 5 | -0.016 | 0.266 | 0.440 | 0.106 | 0.774 | 0.063 |
| Europe & Central Asia | 7 | 0.209 | 0.181 | 0.809 | 0.064 | 0.672 | 0.164 |
| Middle East & North Africa | 2 | -0.281 | 0.455 | 0.140 | 0.149 | 0.161 | 0.247 |
| West & Central Africa | 18 | -0.288 | 0.122 | -0.403 | 0.071 | -0.260 | 0.111 |
| Eastern & Southern Africa | 16 | 0.078 | 0.087 | -0.006 | 0.088 | 0.388 | 0.070 |
| Latin America and Caribbean | 8 | 0.589 | 0.035 | 0.322 | 0.098 | 0.722 | 0.051 |

Note: These analyses used the countries as units of analyses, so they are not affected by the country population size.

## Conclusion

We explored whether a slightly modified version of the original SWPER, previously validated for African countries (10), could be used as an indicator of women’s empowerment across LMICs. 14 items identified from the previous version, that allow the assessment of empowerment in three dimensions (attitude to violence, social independence, and decision-making), were pre-selected for the SWPER Global validation process. The PCA analysis, showed consistency across countries in terms of patterns of components retained and item loadings for the attitude to violence and decision-making domains. For the social independence domain, we found more variability in terms of the item loadings. In particular, women’s education had low loadings in regions with higher educational levels, and therefore less variability in educational levels. Notwithstanding, the correlation between the country-specific and the SWPER Global scores were above 0.89 for all three empowerment domains in all 62 countries included in the analysis.

Two important changes from the previous version of the SWPER were made based on the workshop held with experts on women’s empowerment. The first was to remove the women’s working status as it was agreed that there are many reasons why women work, which are not necessarily related to their own choices. Also, work may not be empowering, depending on the working conditions, and on whether or not women are formally employed, are entitled to maternity leave, are paid for their work, and are able to decide what to do with their earnings. Our preliminary analyses (results not shown) showed that in several countries the work item had negative loadings in the PCA, suggesting that working could be indicative of lower empowerment levels. This inconsistent behavior has also been demonstrated in the literature. For example, women who were working and paid in cash in Egypt were at lower risk of experiencing intimate partner violence, while in India, Peru and Iran their risk was higher (25,26). The second change in the SWPER Global was to give equal weights to women’s involvement in household decisions, whether they were taken only by the women or jointly with her the partner. In the original version of the SWPER, a decision made by the woman alone was given a higher score than a joint decision with the partner. However, there was consensus among experts that joint decisions on household issues such as purchases and visits to family or friends were thought to be a potential reflection of gender equality (27). These content modifications were essential to refine and expand the index to a global context including all LMICs.

Our findings show the SWPER Global is a suitable common measure of women’s empowerment for LMICs, addressing the need for a single consistent survey-based indicator of women’s empowerment that allows to track progress over time and across countries at the individual and country levels. The external validity assessment showed overall strong correlations between country rankings in the SWPER Global with their rankings in the GDI and the GII which capture distinct aspects of gender inequalities at the country level. Construct validity of the SWPER was also previously assessed at the individual level, through its positive association with modern contraceptive use, institutional delivery, stunting (10) and, more recently, with child developmental outcomes (28) and neonatal, infant and under-5 mortality (29). In the current manuscript, although we chose not to present associations between the SWPER Global with health outcomes, additional analyses performed show its positive associations with the coverage of reproductive, maternal, newborn and child health interventions in LMICs (30), reassuring its potential to widen the research on the effects of women’s empowerment on health interventions and outcomes.

After the first version of the SWPER was published in 2017, other similar measures of women’s empowerment in the context of Africa have been proposed in the literature. In 2018, Asaolu et al. (31), using DHS from nineteen countries across Sub-Saharan Africa, identified attitude towards violence, labor force participation, education, and access to health care as valid domains of empowerment although some variations between sub-regions were observed (e.g. in East Africa education was surprisingly not a relevant component of women’s empowerment) (31). Also in 2018, Miedema et al. (16) tested the cross-national invariance of women’s empowerment using DHS data from five countries in East Africa and identified three domains of women’s empowerment capturing women’s human/social assets, attitudes related to wife abuse, and women’s participation decision applicable across countries, very similar to what was presented in the original SWPER paper (10). These other efforts highlight the importance of the topic and how different approaches can lead to different but similar results. Also, how diverse priorities also lead to different approaches. In our case, we compromised on country specificity to obtain an indicator that allows for comparative analysis and monitoring across a large array of countries.

The development of a global common measure of women’s empowerment, however, represents a huge challenge, given its complex and multidimensional constructs as well as their variability across different societies with contextually specific gender vulnerabilities. As a result, the SWPER Global has limitations, some already previously recognized and discussed in our previous publication (32). Empowerment is not primarily an outcome, but a process that includes critical consciousness, aspiration, voice, choice, and change (33,34); and as such, there are elements enabling or limiting it that have not been considered in the SWPER (32). Decision-making assesses control but not choice, nor risk for sanctions or backlash based on choices made. Inclusion of aspects such as personal ownership of assets, economic participation, and opportunities and participation in governance processes were also pointed in the expert workshop as important measures of empowerment that are not captured by the SWPER. Some of these items are not available in DHS surveys and would require different data sources. However, there is a growing number of measures related to assets, opportunity and agency that are being added to DHS with potential to be incorporated in future updates of the SWPER Global. These include mobile phone ownership, internet access, bank accounts, and decision-making on how to spend personal earnings. As these are only available for a few recent surveys, their inclusion in the SWPER at this point would markedly reduce the number of countries for which the index could be calculated. Also, more items are needed to capture additional domains of women’s empowerment, especially in contexts with higher socioeconomic development, including (but not being restricted to) economic empowerment, power relations outside marriage including political engagement or influence, social and occupational leadership and positioning, and freedom of movement and safety at the individual level.

Another important limitation is that the index is limited to women married or in a union and therefore leaves out sizeable groups of women in some countries (Table S4 in the supplementary material). African and Asian countries tend to have higher proportions of married adolescent girls, which may affect comparisons of empowerment levels across world regions and require age-stratified analyses. The development and validation of a measure that includes both partnered and unpartnered women, however, would require a very different approach, which is beyond the scope of the proposed indicator based on national health surveys for which most of the relevant information are only available for women that are married or in a union. Lastly, it is important to note that due to data availability, the countries included in our analysis represent 48% of all LMICs, ranging from over 70% of the countries from South Asia, West and Central Africa and Eastern and Southern Africa to 14% of the countries from Middle East and North Africa (Table S5 in the supplementary material). Given the little difference found in the behavior of the SWPER between the original version for Africa and the current, including all available LMICs, it is unlikely that the inclusion of more countries would lead to relevant changes to the indicator we proposed.

Acknowledging the current limitations of the SWPER Global, particularly in light of the recommendations from the expert workshop, efforts must continue to improve the indicator. Nevertheless, the SWPER Global is a pioneer indicator of women’s empowerment based on individual-level survey data from LMICs, which enables comparability between countries from all world regions and over time. It represents an advance over other global gender and development indices by including a domain on attitudes towards violence against women, a prevalent form of human rights violation as well as an important universally recognized aspects of gender inequality worldwide. Use of the SWPER Global will contribute to monitoring the progress towards SDG5, in terms of gender equality and empowerment at the national and sub-national levels. It also allows quantification of the role of empowerment in terms of health, nutrition and developmental outcomes of women and children worldwide.

## Data Availability

All data relevant to the study are included in the article or uploaded as supplementary information. The data used in this manuscript are publicly available, anonymised, and geographically scrambled to ensure confidentiality. More information on DHS can be found at https://dhsprogram.com/, where survey datasets can be downloaded.

https://www.dropbox.com/sh/cw6e2th24l4ausa/AAC792VQx3CL7CuIdLwTxKiWa?dl=0

## Contributors

FE conducted the analysis and drafted the manuscript. AR, CV, FH and CC contributed to the writing of the manuscript and to the critical revision of the analysis. AB originally proposed the idea of the indicator, advised on the analysis, and contributed to the writing and revision the manuscript. All authors read and approved the final manuscript.

## License

The Corresponding Author has the right to grant on behalf of all authors and does grant on behalf of all authors, an exclusive license on a worldwide basis to the BMJ Publishing Group Ltd to permit this article (if accepted) to be published in BMJ and any other BMJPGL products and sublicenses such use and exploit all subsidiary rights, as set out in our license.

This is an open access article distributed in accordance with the terms of the Creative Commons Attribution (CC BY 4.0) license, which permits others to distribute, remix, adapt and build upon this work, for commercial use, provided the original work is properly cited. See: http://creativecommons.org/licenses/by/4.0.

## Data availability statement

All data relevant to the study are included in the article or uploaded as supplementary information. The data used in this manuscript are publicly available, anonymised, and geographically scrambled to ensure confidentiality. More information on DHS can be found at https://dhsprogram.com/,where survey datasets can be downloaded.

### Acknowledgements

We would like to thank the Pan American Health Organization and the Countdown to 2030 Initiative for co organizing a workshop with experts in women’s empowerment and gender inequalities to discuss the applicability of the SWPER in other world regions and, more specifically, in Latin America and Caribbean countries. We are also very grateful for the valuable contributions of all the participants, whom we wish to acknowledge by name. However, we are still in the process of obtaining the written consent, so the names will be included later on.

## Ethics committee approval

This paper works specifically with Demographic and Health Surveys data, for which the ethical responsibility is entirely of the institutions that conducted the surveys in each country, eliminating the requirement of this study’s ethical approval.

